# Cavernoma in septum pellucidum: descriptive analysis and review of existing literature

**DOI:** 10.1101/2021.02.18.21252024

**Authors:** Samiul Haque, Asraful Islam, Tyfur Rahman, Mohammad D. Hossain, Abu Bakar Siddik, Md Manjurul Islam Shourav, Md Fayad Hasan, Samar Ikram, Masum Rahman

**Author notes:** Authors has equal contribution (Samiul Haque, Asraful Islam). Conflict of interest: All authors declare no conflict of interest. Disclosures: The authors have no financial conflicts of interest to declare. Human/Animal Rights: This article does not contain any studies with human or animal subjects performed by any of the authors.

## Abstract

**Background:** Cavernomas are rare central nervous system (CNS) lesions that constitute a distinct type of vascular malformation encountered in the brain parenchyma or ventricular system. A cavernoma can be familial or sporadic forms and exhibit a range of presentation from incidental findings to seizures, headaches, hemorrhage. Septum pellucidum cavernoma is exceedingly rare and should be studied for its unique topographical location and clinical course.

**Method:** We performed a comprehensive literature search and review using multiple databases. the title/abstract and MeSH keywords used included “cavernoma,” “cavernous hemangioma,” “cavernous malformation,” “cavernous angioma,” “CM,” “septum pellucidum” “SP” and “intraventricular,” along with “AND” and “OR” operators. Demographic and clinical data of each patient were collected for qualitative synthesis.

**Result:** Reported cases were diagnosed at a median age of 42 years; the most frequent symptom was headaches. The incidence of hemorrhage and hydrocephalus was 30%. Gross total resection was performed in 100% of patients and exhibit clinical improvement.

**Conclusion:** The unique location of the cavernoma exhibits clinical presentations seen and the surgical approach used. Gross total resection conveys the impression of optimum management strategy and leads to a magnificent outcome in most cases.

## Introduction

Lesions affecting septum pellucidum are broad and include tumors, cysts, hamartomas, and hemorrhages, which usually arise from the nearby structures. Among these broad differentials, cavernomas malformation (CM) in the septum pellucidum is rare. Cavernoma or cavernous malformation is an incidental or hereditary hamartomatous vascular malformation. They consist of an enlarged, lobulated, and thin-walled vascular malformation without cerebral tissue, typically residing within the brain parenchyma. In the general population, the overall prevalence of cavernomas malformation is 0.02-0.13%, whereas 5-13% among CNS vascular malformations. CNS cavernomas are more frequent in the supratentorial region (74–90%) than infratentorial (10–26%), whereas intraventricular cavernomas are a rare variety of CNS cavernomas accounting for 2.5-10% of all intracranial cavernomas. [1, 2, 3]

The prevalence of a cavernoma in the septum pellucidum (SP) is infrequent, having characteristic symptomatology and radiological features. The unusual location, relevant clinical manifestations, risk factors, radiological characteristics, and management principles of SP cavernomas are summarized following existing literature review. We reviewed relevant literature to find the answer to the following questions: 1) What characteristic clinical features manifest with CMs located at septum pellucidum? 2) What are the associations of septum pellucidum cavernoma? 3) How frequently hydrocephalus, seizure and hemorrhage manifest in patients with these lesions? 4) What is the efficacy of surgical resection in the treatment of these lesions? 5) What surgical approaches have been used when treating CMs at this specific location? 6) What are the outcomes of CM of this location in terms of recurrence, and survival?

### Data Sources and Search Strategies

A comprehensive search of several databases from 1990 to January 15th, 2021, limited to English language and human study, was conducted. The databases included Pubmed, Ovid MEDLINE(R) and Epub Ahead of Print, In-Process & Other Non-Indexed Citations, and Daily, Ovid EMBASE, Ovid Cochrane Central Register of Controlled Trials, Ovid Cochrane Database of Systematic Reviews, and Scopus. The search strategy was designed and conducted by an experienced librarian with input from the study’s principal investigator. Controlled vocabulary supplemented with keywords was used to search for patients with septum pellucidum cavernomas. Keywords used, “septum lucidum,” “septum pellucidum”, “septum pelusidum,” “supracommissural septum,” “Cerebral ventricle,” “neoplas,” “tumor,” “tumour,” “angioma,” “hemangioma,” “haemangioma,” “cavernoma,” “cavernous,” along with “AND” and “OR” operators. PICO question narrated for the study, **P**opulation: any patient with septum pellucidum cavernoma, **I**ntervention: surgical resection, **C**omparator: no comparator, **O**utcomes: symptomatic improvement, survival, recurrence, progression. There were no restrictions imposed on the age, gender, ethnicity of the patient.

Two independent authors screened the search results by examining titles and abstracts for relevance. Articles were finally selected if they met inclusion criteria following full-text article review. Disagreements were resolved by a third reviewer. Any conference papers or studies without full texts, review articles, or meta-analyses without new cases were excluded. Following data were collected from included papers in a preformulated electronic database: age, sex, presenting symptoms, site, and characteristics of the lesion, presence of bleeding, management, prior radiation, presence of concurrent or prior CNS cavernoma, presence of hydrocephalus, management details, surgical approaches and results, clinical outcome, and follow-up. To overcome reporter’s bias, only patient-reported symptomatic status was compared to preoperative condition.

## Results

10 studies were selected using the PRISMA flow diagram and were included in the final analysis. The literature included 10 patients with CMs in the septum pellucidum (Table-1) with a median age of 42 years (range: 16 to 74 years) [4, 3, 5, 6, 7–11]. Of these patients, 8 were male (80%) and 2 were female (20%). Among the study population, the most common presenting symptom was headache (70%, n=7). 20% of patients reported memory impairments. Seizure was reported in one patient precipitated by head trauma (patient-1). Hydrocephalus was apparent on imaging in 30% of cases. 30% had intralesional bleeding at the time of presentation. One asymptomatic patient with prior cavernoma in different locations was diagnosed incidentally on a follow-up MRI. None had a positive family history mentioned in the reviewed literature. In the case of established risk factors of cavernoma formation, one of the patients had a history of prior radiation and another had a history of head trauma. There was insufficient data on other risk factors. The most common finding on MRI of the brain was a heterogeneous contrast-enhancing lesion (n=9), consistent with cavernoma. In 40% of patients (n-4), lesions were limited to septum pellucidum, while 60% (n=5) had extension into either ventricle. All patients managed surgically without radiation, either open procedure (90%) or endoscopic (10%), and the anterior interhemispheric transcallosal approach was used in most of the cases (70%). Gross total resection was achieved in all patients with negligible postoperative complications. Follow-up data were reported in 80% of cases and did not yield any recurrence or progression.

**Table 1:** Patient demography, clinical manifestation and relevance

| No . | Author | Patient s | Presenting symptoms | Family History | Prior or concurrent CNS cavernoma | Prior radiation |
| --- | --- | --- | --- | --- | --- | --- |
| 1 | Paiva A | 19, M | Incidental finding following head injury and seizure | Absent | Absent | CNS Radiotherapy |
| 2 | Baldo S | 32, F | Asymptomatic, diagnosed by follow-up MRI for other CVs | Absent | Multiple, prior | Absent |
| 3 | Entezami P | 16, M | Headache | Absent | Absent | Absent |
| 4 | Faropoulos K | 56, M | Headache and lassitude | Absent | Absent | Absent |
| 5 |  | 31, M | Headache, loss of consciousness, accompanying vomiting | Absent | Absent | Absent |
| 6 | Muzumdar D | 35, F | headaches and giddiness | Absent | Absent | Absent |
| 7 | Katoh M | 44, M | Hemorrhage, Memory disturbance | Absent | Absent | Absent |
| 8 | Kasliwal MK | 55, M | Headache | Absent | Absent | Absent |
| 9 | Constantinos Picolas | 58, M | Headache, personality and memory changes, intraventricular hemorrhage | Absent | Absent | Absent |
| 10 | Motaz Alserehi | 74, M | Headache, vomiting, and unsteady gait for a 2-week duration, intraventricular hemorrhage | Absent | one, in the left putamen. | Absent |

## Discussion

Cavernomas are a cluster of abnormal blood vessels of benign histology. These benign malformations are found in the Central nervous system and throughout the body and have been called Cavernomas, cavernous angiomas, and cavernous hemangiomas. They belong to those cerebrovascular malformations that have no arteriovenous shunt and thus are generally angiographically occult [12]. These malformations are sporadic or occur in relation to an inherited pattern [4]. They are commonly anticipated in the supratentorial region than the infratentorial region exhibiting a varying prevalence of approximately 70-80% and 10-26% respectively [13–15]. Intraventricular cavernomas are rare, commonly found in the lateral ventricle followed by the third ventricle and the fourth ventricle in the CNS.

The septum pellucidum (Latin for “translucent wall”) is a thin, triangular, vertical double membrane separating the anterior horns of the left and right lateral ventricles of the brain. The first reported case of cavernoma in the septum pellucidum available in the English literature was an intraventricular hemorrhage and the patient presented with headache, nausea [16]. The lesion was resected with no residual signs and symptoms.

### Clinical Presentation

Symptoms encircling cavernoma are supposedly induced by a mass effect, raising the intracranial pressure manifesting as headache, nausea, vomiting, papilloedema. CNS cavernomas present with a varying spectrum of clinical signs and symptoms depending on the anatomical site. In the supratentorial region, they present with headaches, seizures, and focal neurological deficits. The severity of such clinical features depends on the location and the extent of the size of the entity. Seizure is a common presentation in supratentorial cavernoma, approximately 39-79% of patients with an annual risk of development of 1.5% [7,17,18] In contrast, bleeding is more common in the infratentorial region with a prevalence of 3.8% compared to the supratentorial region, which bears a prevalence of 0.4%. Our analysis demonstrates hemorrhagic complications in 30% and seizure manifestation in 10% of cases, which is quite the opposite of the statistics of supratentorial cavernoma. The manifestation of CNS cavernoma may be delayed, depending on their size and location. Even though relatively small lesions and having enough room on both sides in lateral ventricles, 90% of our patients were symptomatic on presentation. We hypothesize minute intralesional bleeding for symptomatic manifestations. These fragile vascular malformations are susceptible to hemorrhages and are eventually surrounded by hemosiderin deposits and a gliotic membrane when they encounter clinically silent bleeding or may have hemorrhagic stroke-like features.

Anatomically septum pellucidum runs as a thin sheet from the corpus callosum down to the fornix. The fornix plays an important role in memory, connecting the hippocampus and other structures involved in memory, including the anterior thalami, septal nuclei, and the mamillary bodies [8,19]. An impact on the hippocampus can affect the ability to form long-term memories leading to anterograde amnesia.

One study has shown, memory disturbances in a patient with intraventricular cavernoma are more likely (57.1%) when affecting the lateral wall of the third ventricle, mostly secondary to compression on columns of the fornix running through it. [20,21] A confined hemorrhage within septum pellucidum cavernoma may damage the bilateral fornices. Therefore, even with a small lesion, septum pellucidum cavernoma should undergo immediate surgical removal without further haste. CNS cavernoma may also cause obstructive hydrocephalus when interrupt CSF flow. It has shown that hydrocephalus is more prevalent while affecting the foramen of Monro.[20] In our analysis, hydrocephalus was found in 30% of reported cases of septum pellucidum cavernoma, relatively lower than third ventricular cavernoma (59%). Hydrocephalus likely occurred in these patients owing to downwards extension of the lesion compressing foramen of Monro.

### Imaging

Intraventricular lesions exhibit several differential diagnoses, including arteriovenous malformations, astrocytomas, choroid plexus papillomas, ependymomas, and colloid cysts [4,6]. Radiologically, cavernoma have been typically described as “popcorn” or “berry” shaped appearance on MRI which represents a histologic pattern of fibrosis, thrombosis, calcification found within these lesions. MRI is the gold standard for imaging cavernomas. Cavernomas are angiographically occult malformations since they are not connected to any major blood vessels [22]. Nevertheless, developmental venous anomalies typically associated with and are visible during angiography and MRI [4]. In the parenchymal form, a hypointense ring is observed on T1 and T2-weighted images due to hemosiderin deposits from recurring microhemorrhages [4]. In our analysis, all cases exhibit characteristic features on MRI includes, well-circumscribed nodular lesion with heterogeneous signal intensity, none to moderate post-gadolinium enhancement with or without hemorrhagic components. (Table-2)

**Table 2:**
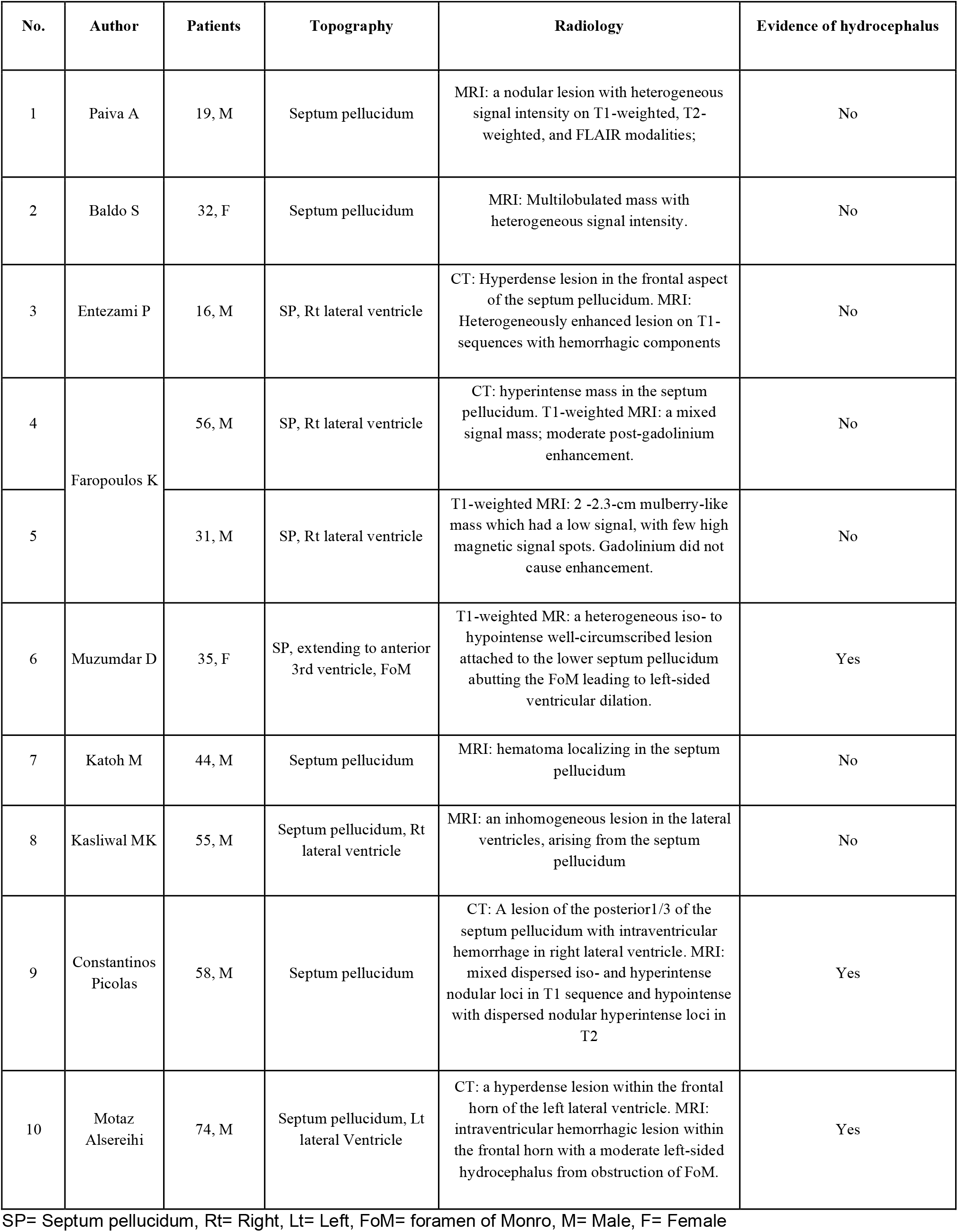
Imaging data

### Management

The management of intracerebral cavernomas is related to specific locations and signs and symptoms. Asymptomatic lesions in eloquent areas can be observed. Whereas complete excision is the treatment of choice symptomatic patients with mass effect (headache and hydrocephalus, neuro deficit), seizures, recurrent hemorrhages, intractable seizures, and lesions increasing in size. [3,7,20]

Intraventricular cavernomas are usually accessed by transcranial, transcortical approach, which is technically easier requires large ventricles and a craniotomy to enter the brain. Like all procedures, there are potential risks to be discussed as the judgment of benefits versus risk must be considered to proceed with the procedure. In the end, the patient benefit must outweigh the risks. A transcranial transcortical approach can cause epilepsy. The other approach is a transcranial interhemispheric transcallosal route. The anterior transcallosal approach is adequate and is often superior to the ventricular transcallosal approach without hydrocephalus (to prevent blood spillage into the ventricles to prevent meningism and obstructive hydrocephalus. This approach is more anatomical and does not require a large ventricle, but it is technically more demanding.[3] In addition, callosotomy is too posterior and can give away a disconnection syndrome problem. An alternative and much less invasive procedure concerns the endoscopic method, which is increasingly used in the treatment of ventricular tumors. Likewise, it brings about technical dismay and hesitation among surgeons due to the vascular nature of these lesions and possible difficulty in achieving hemostasis. [3] Regardless, minimally invasive surgery permits a better surgical outcome and reduces adverse outcomes and prompts better postoperative recovery.

Neuroendoscopy is being increasingly used in the treatment of intraventricular tumors, however its potential for use in the excision of septum pellucidum tumor is still unclear. Since an endoscopic approach leaves out the doubt of achieving adequate hemostasis, microsurgical approach is the viable option to ensure a safe and complete resection, given the anatomy of the region and its intense vascularization [7]. One patient in our cohort (patient-2) underwent endoscopic resection, which was justified by the possibility of heavy scar from previous surgery. Inferior parietal transcortical approach was reported in patient-11 due to presence of ventricular hemorrhage [11].

To summarize the approach and risks. For a lateral ventricle trigonum IVC, transtemporal, posterior transcortical or posterior transcallosal approach is advocated [23–25]. And likewise, each approach bears its own risk. The posterior middle temporal gyrus transcortical approach can induce damage to the optic radiation leading to left lower quadrantanopia or to the dominant hemisphere. Posterior parietal transcortical access can inflict visuospatial apraxia if the mass is located in the dominant hemisphere [6,26]. Ultimately, in the long run, complete surgical removal of IVC is the treatment of choice with low-rate complications and associated morbidity [6]

Radiotherapy is controversial for managing intraventricular cavernoma. Intraparenchymal cavernomas in high-risk areas have been reported to treat with stereotactic radiotherapy [27,28]. In few case reports, they received radiation upon misdiagnosis as a tumor and have not shown satisfactory clinical improvement [29,30]. Moreover, radiation contains the potential risk of bleeding, recurrence, and expansion of the cavernoma. One study has shown that surgical resection of supratentorial cavernomas results in a lower incidence of rebleeding and better control seizures than stereotactic radiotherapy [27]. In our analysis, gross total resection was achieved in all patients with favorable postoperative and long-term outcomes without any radiotherapy.

### Effect of Resection on Outcome

Our analysis denotes gross total resection is an effective method of treating septum pellucidum cavernoma, as 100% of cases demonstrated significant clinical improvement after surgery (Table-3). Nevertheless, memory disturbances were the most common residual deficit following surgery that was persistent in all patients who initially had. We hypothesize irreversible damage to fornices either due to mass effect or minute bleeding. This signifies the necessity of early intervention to treat SP cavernoma, even in asymptomatic cases.

**Table 3:** Management and outcome

| No. | Author | Patients | Surgical Approach | Extent of resection | Histopathology | Post-operative course | Recurrence |
| --- | --- | --- | --- | --- | --- | --- | --- |
| 1 | Paiva A | 19, M | Anterior interhemispheric transcallosal | Gross total resection | NA | Uneventful, no seizure recurrence | NA |
| 2 | Baldo S | 32, F | Transcortical endoscopic | Gross total resection | Consistent with cavernoma | Uneventful, | No recurrence in 5years |
| 3 | Entezami P | 16, M | Anterior interhemispheric transcallosal | Gross total resection | Consistent with cavernoma | Uneventful, headache resolved | No recurrence in 5years |
| 4 | Faropoulos K | 56, M | Right frontal craniotomy* | Gross total resection | NA | Uneventful | NA |
| 5 |  | 31, M | Frontotemporal craniotomy* | Gross total resection | NA | Uneventful | No recurrence on repeat MRI |
| 6 | Muzumdar D | 35, F | Anterior interhemispheric transcallosal | Gross total resection | Consistent with cavernoma | Uneventful, headache resolved | No recurrence in 5years |
| 7 | Katoh M | 44, M | Anterior interhemispheric transcallosal | Gross total resection | Consistent with cavernoma and prior hemorrhage | Uneventful, memory unchanged | NA |
| 8 | Kasliwal MK | 55, M | A right frontal transcortical approach | Gross total resection | NA | Uneventful | NA |
| 9 | Constantinos Picolis | 58, M | Inferior parietal transcortical trans-ventricular | Gross total resection | Consistent with cavernoma | Uneventful, Mild short-term memory disturbances persisted | No recurrence in 6 months |
| 10 | Motaz Alserehi | 74, M | Anterior interhemispheric transcallosal | Gross total resection | Consistent with cavernoma and concurrent presence of subependymoma | Uneventful with symptomatic improvement | No recurrence in 1 year |

NA= Not available, \*author's assumption of anterior interhemispheric transcallosal according to the most likely approach with this craniotomy for this lesion.

### Study limitations

It’s a review article that summarizes all previously reported cases without having any primary patient data. The patient sample size is too small to bear any firm conclusion regarding intervention and outcome.

## Conclusion

While common in the parenchyma and ventricle, SP cavernoma is a rare CNS vascular malformation with distinct clinical behavior due to its location. Perilesional lateral ventricular space may allow expansile growth and probably higher hemorrhagic incidences with potential damage to nearby noble structures. Despite smaller lesions, high clinical and radiological suspicion is necessary to expedite early intervention to prevent irreversible neurologic impairment. Complete excision with the long-term excellent and curative outcome is possible and is the treatment of choice.

**Figure 1:**
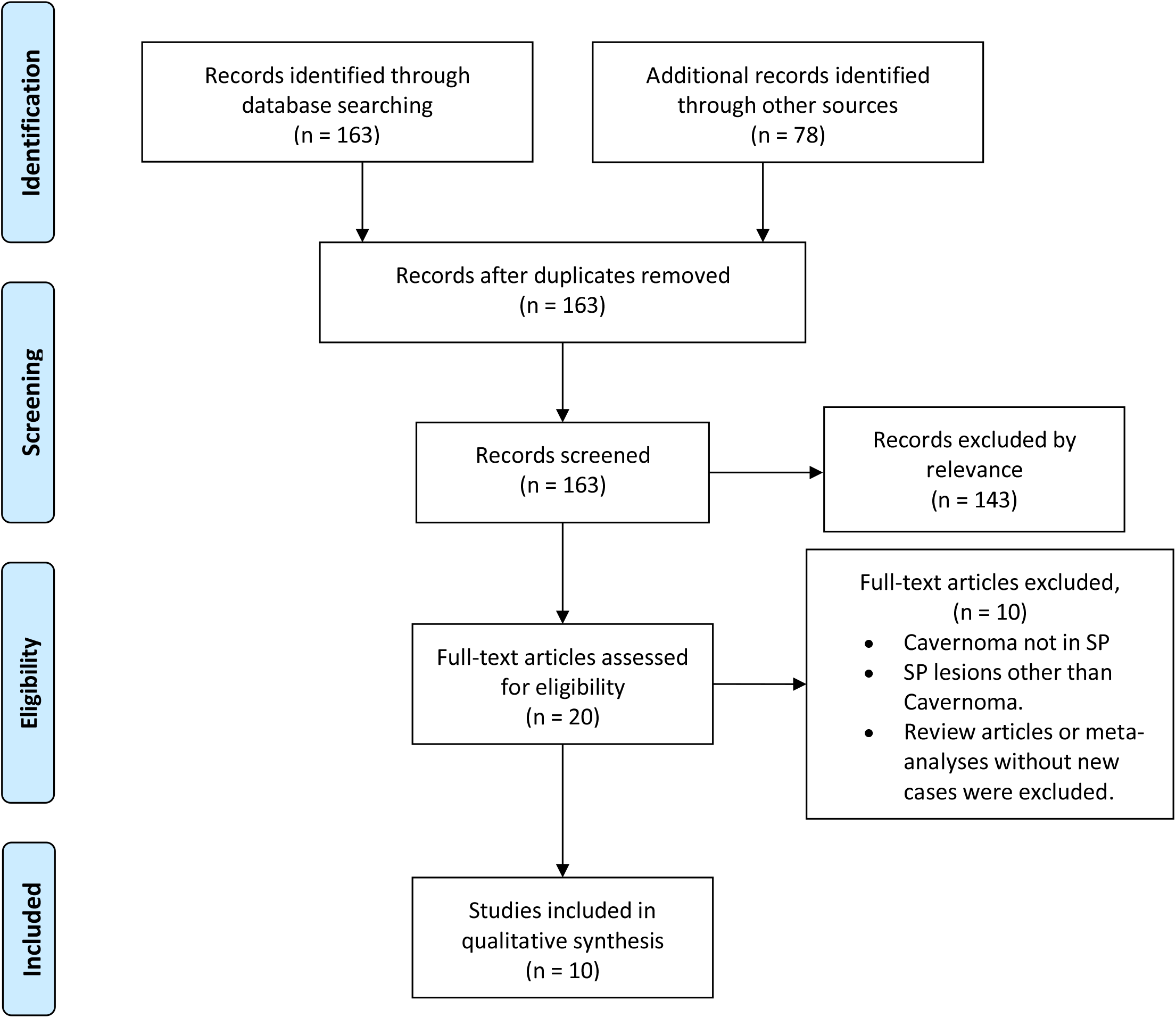
PRISMA flow diagram illustrating the selection process for the studies included in the review for final analysis.

**Figure 2:**
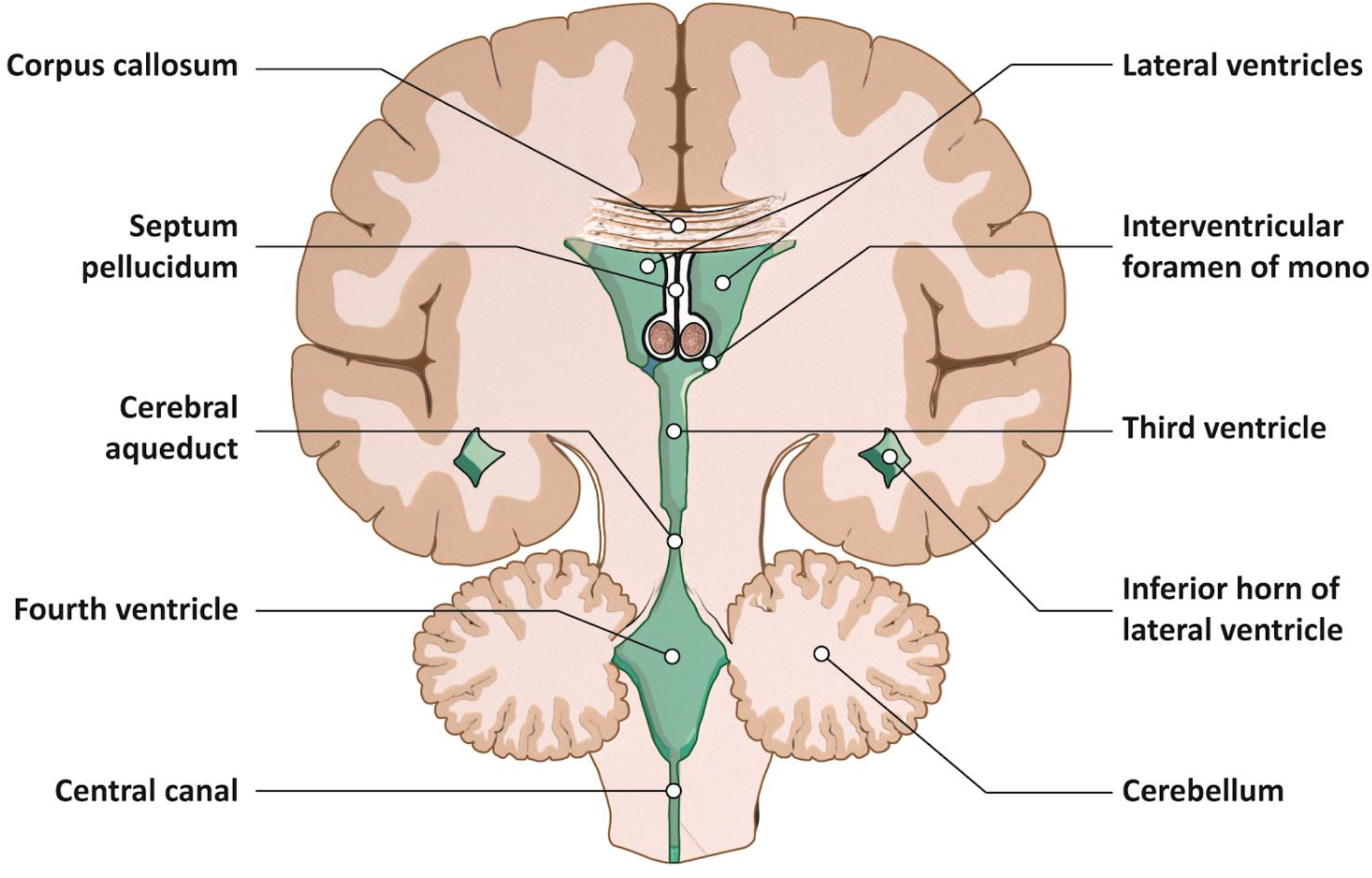
Coronal view ventricles of the brain showing the anatomical location of septum pellucidum.

**Figure 3:**
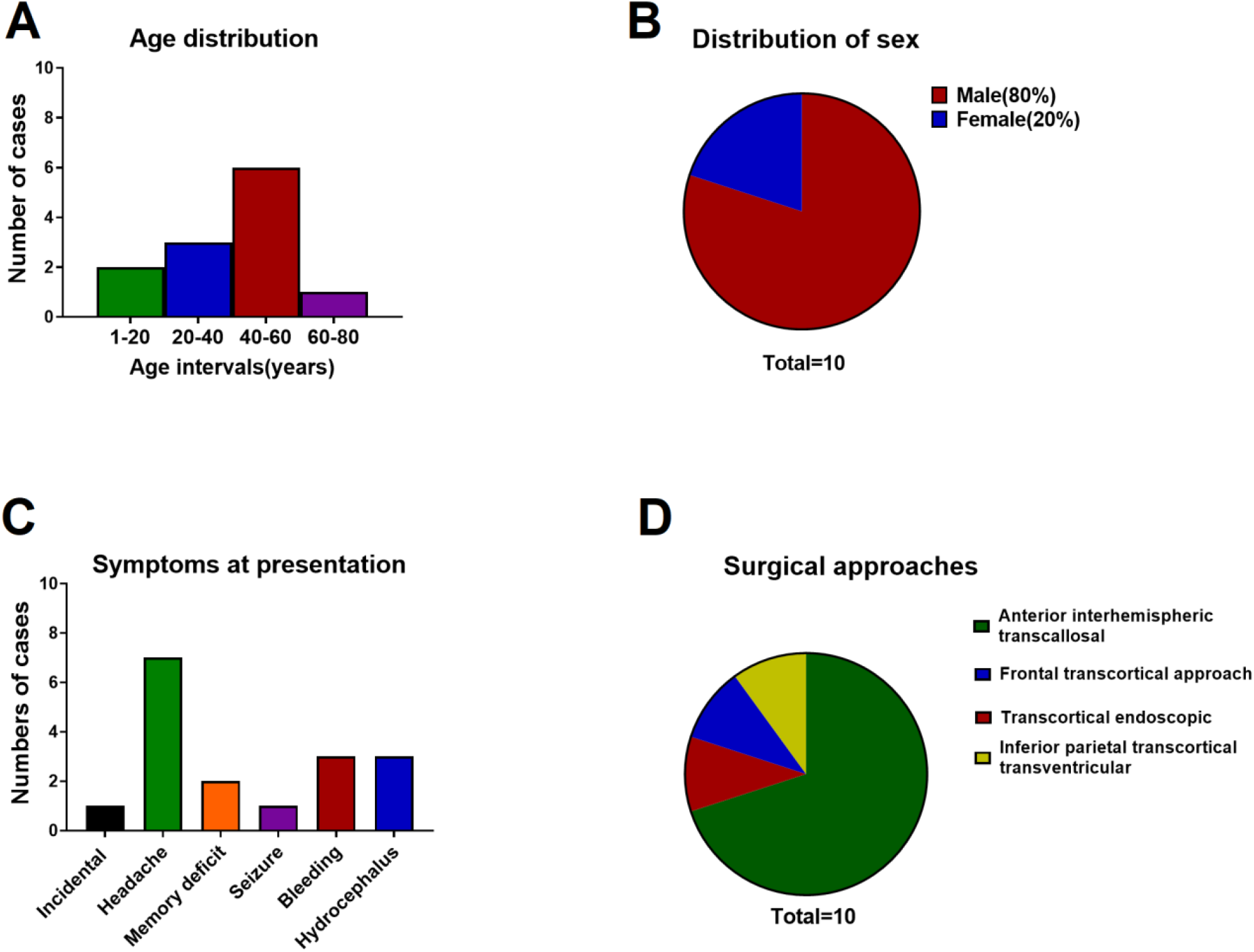
Graphical representation of patient demography, presenting symptoms and surgical approach. A-B: age and sex distribution. C: Frequency distribution of presenting clinical manifestation. D: Frequency of various approaches used for surgical resection.

## Data Availability

Data are available in the main manuscript in the table and diagram.

## Author’s contribution

Literature search, screening, and data extraction by SH, AI, TR. Data analysis and manuscript writing by MDH, AS, MIS, SI. Conceptualisation, study design, data analysis, and manuscript writing and editing by MR. Illustration by MFH. All authors reviewed and suggested edits to make the final version.

## Acknowledgments

We want to acknowledge and thank Larry J. Prokop, Outreach Librarian, Mayo Clinic Libraries, and CMSR (center for medical study and research) foundation.

